# Simultaneous quantification of multiple immune checkpoint interactions in melanoma

**DOI:** 10.1101/2025.07.04.25330865

**Authors:** Cristina Cacho-Navas, Laura Camacho, Jon Agüero, Baterdene Batmunkh, José María Gracia, Carlos Eduardo de Andrea, Salvador Martin Algarra, Markel Rementeria, James Miles, Juan Gumuzio, Fernando Aguirre, Peter J. Parker, Véronique Calleja

## Abstract

Immune checkpoint inhibitors (ICIs) have transformed the therapeutic landscape of advanced malignancies. However, only a subset of patients respond to treatment. Recent efforts have focused on the identification of novel biomarkers that capture the dynamic and functional state of the tumour immune microenvironment, yet their clinical translation has remained challenging. To address these limitations, we applied amplified FRET-FLIM (QF-Pro®) technology to spatially resolve and quantitatively assess functional immune checkpoint interactions directly in cells and FFPE tissue and tumour samples. Using this approach, we demonstrated that PD-1/PD-L1, CTLA-4/CD80, TIGIT/CD155, and LAG-3/MHC-II interactions can be robustly quantified in routine patient samples from multiple tumour types. Co-analysis of all four immune checkpoints was performed within an ICI-treated melanoma patient cohort enabling the identification of patterns of concomitant engagement. Notably, patients with high PD-1/PD-L1 interaction levels typically also exhibited a significantly elevated CTLA-4/CD80 interaction status. Furthermore, the survival analysis of the patients treated with immune checkpoint blockade showed that the LAG-3/MHC-II interaction state discriminated responders from non-responders, independently of the specific ICI therapy administered, suggesting that the assessment of LAG-3 engagement may serve as a novel prognostic biomarker in immunotherapy treatment in melanoma.

## Introduction

Immune checkpoints (ICP) are essential regulatory pathways that maintain immune homeostasis and prevent excessive immune activation [1] (Fig.1A). In cancer, tumour cells evade immune surveillance by triggering ICP engagement [2, 3] and while immune checkpoint blockade against PD-1/PD-L1 (nivolumab, pembrolizumab) or CTLA-4 (ipilimumab), has shown remarkable benefits [4], some patients fail to complete respond to treatment and often suffer strong immune related adverse events (irAEs) [5, 6]. This is particularly important in combination treatments like anti-PD-1 with anti-CTLA-4 [7] or the newly approved anti-LAG-3 (relatlimab) in metastatic melanoma, where the improved response is accompanied by about 60% and 20% respectively of grade 3-4 side effects [8]. Therefore, it is critical to identify the patients who will respond to treatment. To do so, companion and complementary diagnostics tests are presently used. They encompass PD-1 and PD-L1 expression, tumour mutational burden (TMB) or T cell infiltration among others [9]. However, patient stratification based on these biomarkers alone has remained suboptimal, underscoring the need for more robust predictive markers. Although novel biomarkers are under investigation [10], none directly assess checkpoint engagement, the primary target of immune checkpoint blockade.

**Figure 1.**
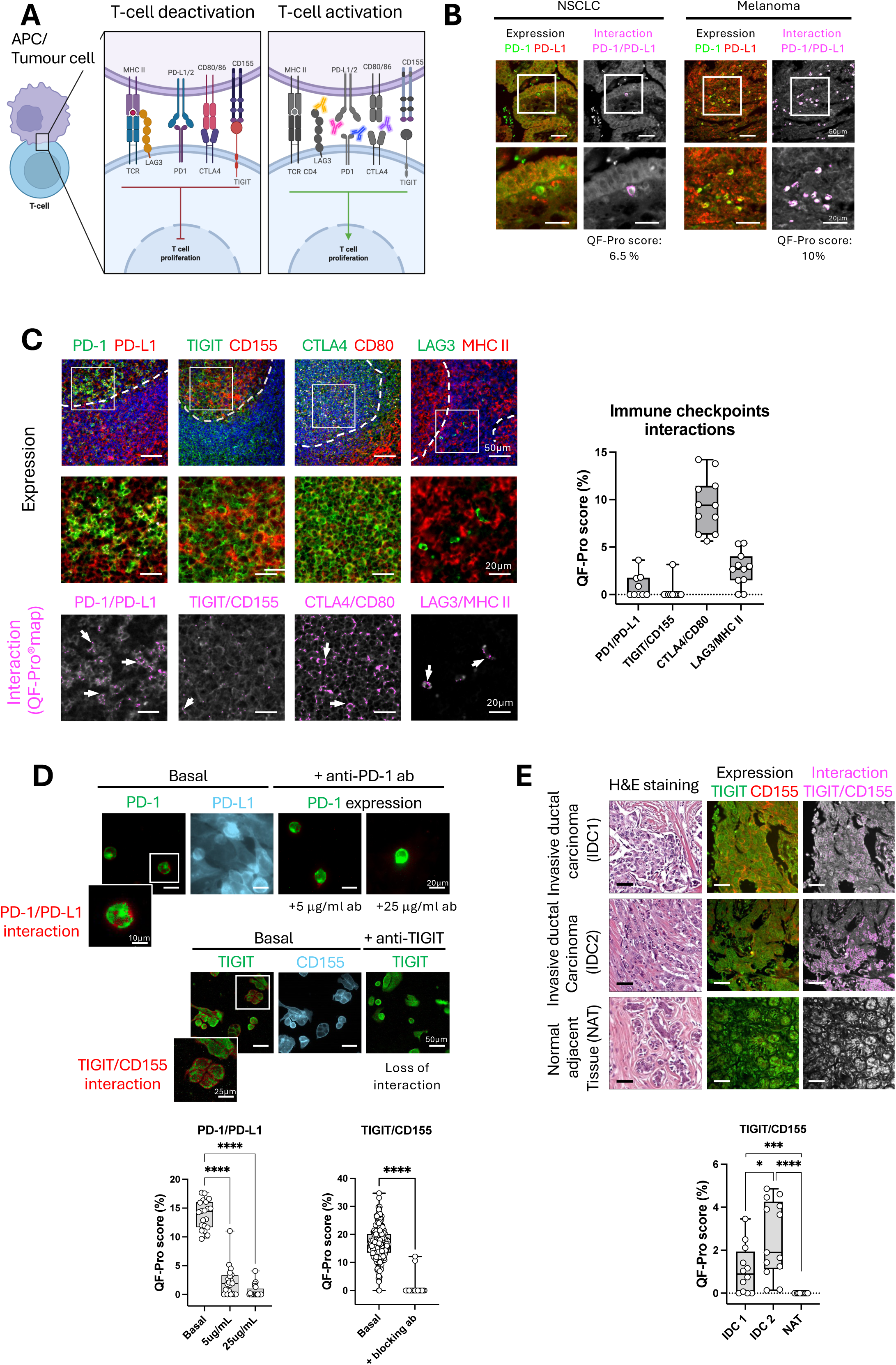
Quantitative measurement of immune checkpoint complex engagement across multiple models. **(A)** Schematic representation of immune checkpoint receptor–ligand complexes and their corresponding therapeutic monoclonal antibodies. **(B)** PD-1/PD-L1 expression (green and red) and interaction (magenta) in NSCLC and melanoma tissues. Interaction maps are overlaid with PD-1 expression (grey), detectable on infiltrating immune cells in both tumour types. ****P ≤ 0.0001. **(C)** Human FFPE tonsil tissue labelled to quantify four immune checkpoint interactions as indicated. Donor (green), acceptor (red), and nuclei (blue) signals are shown for each checkpoint pair. Spatial localisation of immune checkpoint engagement (QF-Pro maps, magenta) is overlaid with donor expression (grey). Box-and-whisker plots display quantitative interaction scores (QF-Pro® scores), with each dot corresponding to a region of interest. **(D)** Validation of PD-1/PD-L1 and TIGIT/CD155 interactions in cell-based co-culture assays using Promega kits, including blocking antibody controls. Biomarker expression is shown in green or blue as indicated, while interactions are shown in red and quantified in box-and-whisker plots, with each dot representing a single cell. **(E)** Invasive ductal carcinoma and adjacent tissues labelled for TIGIT/CD155. TIGIT (green), CD155 (red), and nuclei (blue) expression are shown, with TIGIT/CD155 interaction localization (QF-Pro® map, magenta) overlaid on donor expression (grey).

Biomarkers assessing the functional state of ICP would be expected to better reflect the actual immune status of the tumour microenvironment (TME) and the potential benefit of a specific intervention. This was previously addressed for PD-1/PD-L1 in non-small cell lung cancer (NSCLC), where it was found that the stratification power associated with PD-1/PD-L1 engagement status in patient biopsies showed a far superior predictive value to ICP treatment, compared to the routinely employed PD-L1 TPS/CPS score [11]. These findings prompt the analysis of a broader panel of ICPs, both to understand the patterns of exclusive or coincident engagement elicited by tumours and to provide a framework in which to establish a molecular pathology exploitable in the clinic. Here, we show that multiple checkpoints can be quantified in cells and in FFPE tissue samples of various tumour types (NSCLC, Breast and Melanoma). Moreover the simultaneous determination of PD-1/PD-L1, CTLA-4/CD80, TIGIT/CD155 and LAG-3/MHC-II immune checkpoint axes offered valuable information on ICP co-regulation in in a proof-of-principle study on melanoma patients, potentially guiding the selection of personalised combination regimens and identifying potential biomarkers of response to ICI treatment in melanoma.

## Results

Here, QF-Pro® an amplified FRET-FLIM technology was employed to assess whether in situ engagement of key immune checkpoints (PD-1/PD-L1, CTLA-4/CD80, TIGIT/CD155 and LAG-3/MHC-II) could be measured across cells and multiple tissue types, with the potential to generate clinically relevant insights for immune checkpoint inhibitor therapy (Fig. 1A). The amplified FRET signal enables highly accurate quantification of proteins functional states within complex biological microenvironments, including FFPE tumour samples (Supplementary Fig. 1, Materials and Methods). Primary commercial antibodies to these checkpoints were selected based on (i) epitope recognition within the extracellular domains of the corresponding ligands or receptors, and (ii) prior validation by the suppliers for use in FFPE samples (Supplementary Table 1).

### PD-1/PD-L1 immune checkpoint interaction can be detected in infiltrated immune cells in NSCLC and melanoma tissue

We previously exemplified the predictive value of monitoring PD-1/PD-L1 interactions in assessing response to immune checkpoint inhibitor (ICI) therapy in NSCLC [11]. Associated with the work to extend this approach to a broader collection of checkpoints, we first developed a more comprehensive spatial mapping tool to improve interpretation of findings.

The spatial localization of PD-1/PD-L1 interactions was examined in NSCLC and melanoma tumour tissues, providing additional contextual insight into these interactions. QF-Pro® maps (magenta) revealed that PD-1/PD-L1 interactions occur on infiltrating lymphocytes within the tumor microenvironment (TME) in both tissue types (Fig. 1B). Interaction levels were quantified using QF-Pro® scores, calculated from signals within a defined region of interest (ROI), indicated by the white square and corresponding zoomed image.

### Immune checkpoint interaction detection in FFPE tonsil tissue

With a mapping tool in hand we first sought to validate the four ICP interactions in a tissue model. The engagement of PD-1/PD-L1, CTLA-4/CD80, TIGIT/CD155, and LAG-3/MHC-II was therefore measured in FFPE tissue sections of human tonsil tissue (Fig. 1C). As a secondary lymphoid organ, the tonsil plays a critical role in early immune priming and immune responses, where multiple immune cell populations interact closely with antigen-presenting cells [12]. These characteristics make tonsil tissue an effective model for assessing immune checkpoint interactions in situ.

Interaction quantification (QF-Pro® scores) and spatial localisation (QF-Pro® maps; magenta) were successfully determined for three of the four immune checkpoints. Notably, the TIGIT/CD155 interaction was not detectable in tonsil tissue (one outlier), as evidenced by both the interaction map image and the box-and-whisker plot (Fig. 1C righthand panel). The expression of the corresponding immune receptors and ligands (green and red) was however detected in DAPI-stained tissue sections for all the checkpoints. The data revealed variability in interaction levels as well as distinct spatial localisation patterns among the different immune checkpoints within the same tissue. Importantly, the specific interaction patterns (or lack thereof) observed in the interaction maps are distinct from simple colocalisation of checkpoint protein expression (green and red), supporting the importance of direct interaction mapping. This is particularly evident in the labelling of TIGIT/CD155 where robust expression of both proteins is observed within the same region (see the zoomed middle panel of the boxed area), yet no interaction is visible on the QF-Pro® map (lower panel).

### PD-1/PD-L1 and TIGIT/CD155 interactions measured by QF-Pro® are validated in co-culture models

As all immune checkpoint pairs except TIGIT/CD155 showed detectable engagement in tonsil tissue, we sought to validate the TIGIT complex readout using a cell line model. To this end we adapted the Promega Blockade Bioassay kits to enable measurement TIGIT/CD155 and as a positive control PD-1/PD-L1 interactions using the QF-Pro® platform (see Materials and Methods). These ICP interactions were assessed in co-cultures of CHO-K1 cells engineered to express either human PD-L1 or CD155 and Jurkat cells stably expressing human PD-1 or TIGIT (Promega) (Fig. 1D). PD-1/PD-L1 and TIGIT/CD155 interactions (red) were detected in overlay images with donor protein expression (PD-1 and TIGIT, as indicated shown in green). Notably, these interactions were abolished upon addition of blocking antibodies against PD-1 or TIGIT (respectively) prior to fixation. Quantification of interaction signals and their loss following interfering antibody-mediated blockade is shown in the box-and-whisker plots (Fig. 1D lower panels), demonstrating the specificity of these immune checkpoint engagements in engineered in-cell systems.

### TIGIT/CD155 interaction is observed in invasive ductal carcinoma tissue

Because the interaction between TIGIT and CD155 was detectable in the cell co-culture system, despite not being observed in tonsil, we anticipated that TIGIT/CD155 interaction would be detectable in a tissue where it is more highly enriched. In invasive breast cancer (invasive ductal carcinoma, IDC), upregulation of TIGIT expression has been associated with poor prognosis, suggesting a prominent role for TIGIT/CD155 in the negative regulation of immune responses within the TME in these tumours [13]. Therefore, TIGIT/CD155 interaction was assessed in two IDC samples and normal adjacent tissue (NAT) from a tumour microarray (TMA) (Fig. 1E). Quantifiable TIGIT/CD155 interaction was detected with variability in the tumour samples but no interaction was detected in the non-tumoral tissue (interation maps and box-and-whisker plot). These findings indicate that the TIGIT/CD155 complex can be quantified in IDC, providing direct information on the status of this interaction within these tumours.

### Analysis of ICP interactions in a cohort of advanced melanoma patients treated with immune checkpoint inhibitors as second-line treatment

Having validated the ability of QF-Pro® and the selected antibody pairs to detect PD-1/PD-L1, CTLA-4/CD80, TIGIT/CD155, and LAG-3/MHC-II immune checkpoint interactions in FFPE tissue and tumour samples, we next investigated whether their combined analysis within individual patient samples could reveal patterns of checkpoint engagement. Specifically, the parallel assessment of ICPs interaction, was performed to determine whether these patways are activated concomitantly or operate in a mutually exclusive manner within tumours. To address this we conducted a study in a cohort of melanoma patients (n = 30).

An overview of the clinical characteristics of the melanoma cohort is provided in Supplementary Figure 2 and Supplementary Table 2. The patients received a range of immune checkpoint inhibitor regimens: (i) anti–PD-1 monotherapy (pembrolizumab or nivolumab; n = 8), (ii) anti–CTLA-4 monotherapy (ipilimumab; n = 14), and (iii) combined anti–PD-1 and anti–CTLA-4 therapy (nivolumab plus ipilimumab; n = 8). A forest plot analysis was conducted to evaluate the impact of the clinical characteristics on OS (Supplementary Fig. 2A). Hazard ratio (HR) analysis indicated no statistically significant differences in death rates across the clinical subgroups. However, a trend was observed in which patients with metastatic disease exhibited a reduced median OS compared with non-metastatic patients (21 vs. 32 months, respectively; Supplementary Fig. 2B). Consistent with this finding, metastatic patients also showed a lower rate of clinical benefit at 12 months compared with those with primary, non-metastatic tumours (28% vs. 42%, respectively; Supplementary Fig. 2C). Overall, these analyses suggest that none of the evaluated clinical variables introduced a major bias that could substantially influence the observed results.

### Spatial quantification of immune checkpoints interactions in the ICI-treated melanoma cohort

LAG-3/MHC-II, CTLA-4/CD80, TIGIT/CD155, and PD-1/PD-L1 interactions were quantified and imaged within the tumour regions identified by H&E staining in the melanoma cohort (defined by the yellow outline; Fig. 2A-D) by an experienced pathologist. Representative images were chosen to illustrate patients’ samples with higher ICP interaction versus patients’ samples with lower interaction for each of the four checkpoints. Intensity images show the expression of immune checkpoint receptors (LAG-3, CTLA-4, TIGIT, and PD-1; green) and their corresponding ligands (MHC-II, CD80, CD155, and PD-L1; red) in the regions of interest (ROI) shown. QF-Pro® maps (magenta) depict the spatial localisation of each ICP interaction within the tumour ROI. The distribution of QF-Pro® scores measured at each ROI (individual dots) are presented in the associated box-and-whisker plots, highlighting the interaction heterogeneity and strong statistical difference between these patient examples (P<0.0001). The distribution of the immune checkpoint interactions (QF-Pro® scores) across the melanoma patient cohort is presented on the violin plots (below the images), illustrating further the intra- and inter-patient heterogeneity of immune checkpoint interactions.

**Figure 2.**
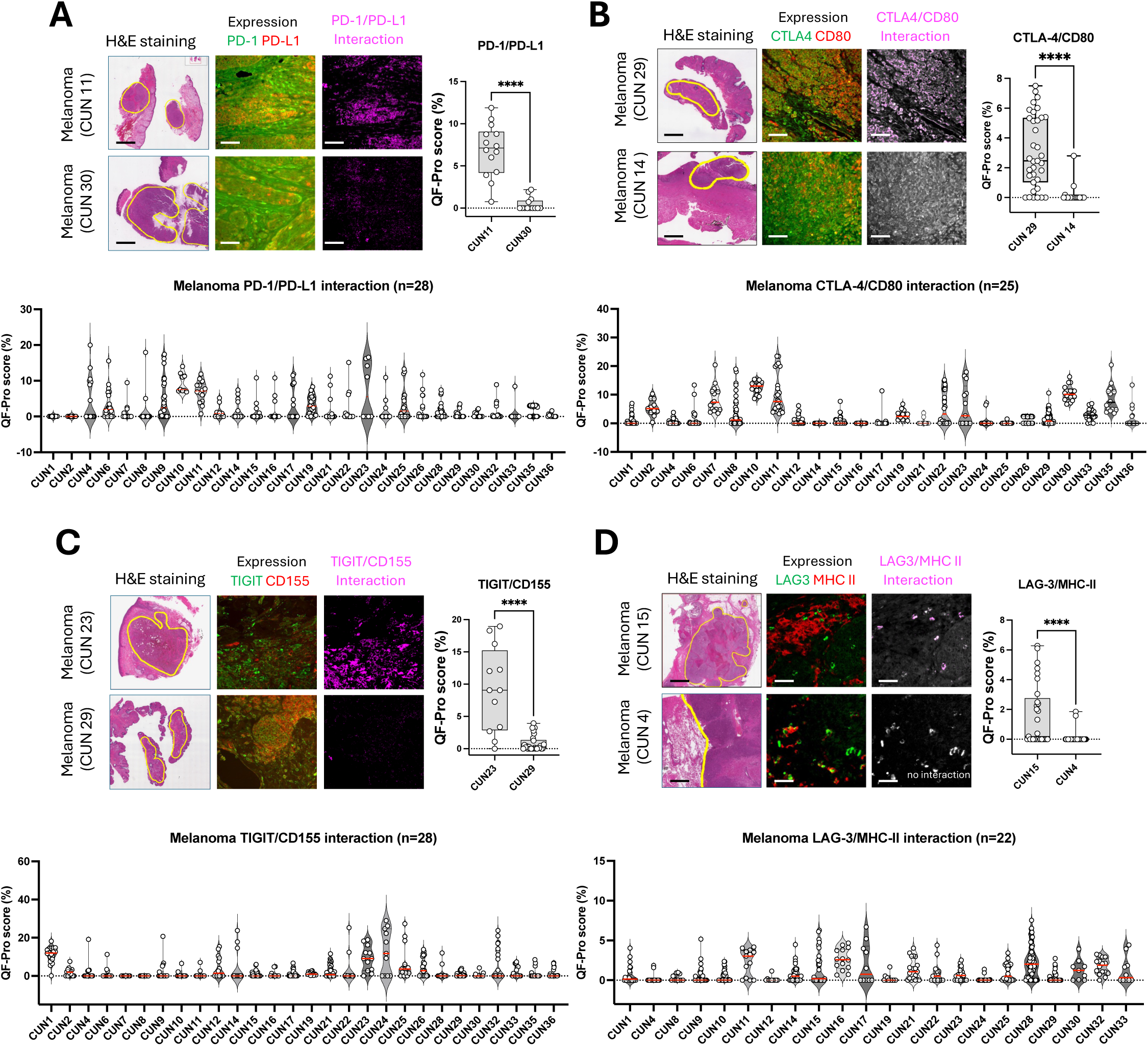
Spatial analysis of ICP engagement in the TME and heterogeneity across the melanoma cohort. **(A–D)** Spatial expression (green and red) and interaction (magenta) of immune checkpoint receptors and ligands are detected using QF-Pro® mapping. Representative patient samples illustrate PD-1/PD-L1 (A), CTLA-4/CD80 (B), TIGIT/CD155 (C), and LAG-3/MHC-II (D) interactions localisation and heterogeneity. The images of the ICP were all taken from the delineate tumour regions (yellow; H&E images). Box-and-whisker plots depict variability in interaction across multiple fields of interest for the same patient, with each dot representing one field of view. ****P ≤ 0.0001. Violin plots presents interaction heterogeneity (QF-Pro® scores) across all patients for the four checkpoints. Each dot represent a region of interest acquired for each patient sample. The median QF-Pro® interaction scores are indicated in red bars.

Notably, analysis of the LAG-3/MHC-II localisation in the TME (Fig. 2D) shows that while both the patients shown present comparable expression of LAG-3 in infiltrated immune cells (see expression panels; green), only patient CUN15 shows interaction (upper interaction panel; magenta). This data highlights further the importance of interaction quantification over protein co-localisation, even when proteins are present within the same spatially defined regions.

### Co-engagement of immune checkpoints

The determination of the interaction status of the four ICPs in each patient within this cohort, offered the unique opportunity to analyse whether the concurrent quantification of the four immune checkpoint interactions in the same patient’s samples could inform on co-regulation or mutual exclusivity of the ICP axis engagement in the tumours.

A Pearsons’ correlation analysis, visualized using a correlation matrix was performed (Fig. 3A). Pairwise relationships between the ICP interaction states were assessed by the correlation coefficients (r) shown in the matrix. R ranges from −1 (perfect inverse correlation; in red) to +1 (perfect positive correlation; in blue), with values close to 0 indicating no linear relationship and with the increasing colour intensity corresponding to stronger associations. The data revealed a positive linear correlation between PD-1/PD-L1 and CTLA-4/CD80. The moderate value of the Pearsons r (r = 0.41; *P* = 0.04) suggested a partial but significant co-engagement of these checkpoint interactions. No other correlations could be observed between other checkpoints combinations within this cohort.

**Figure 3:**
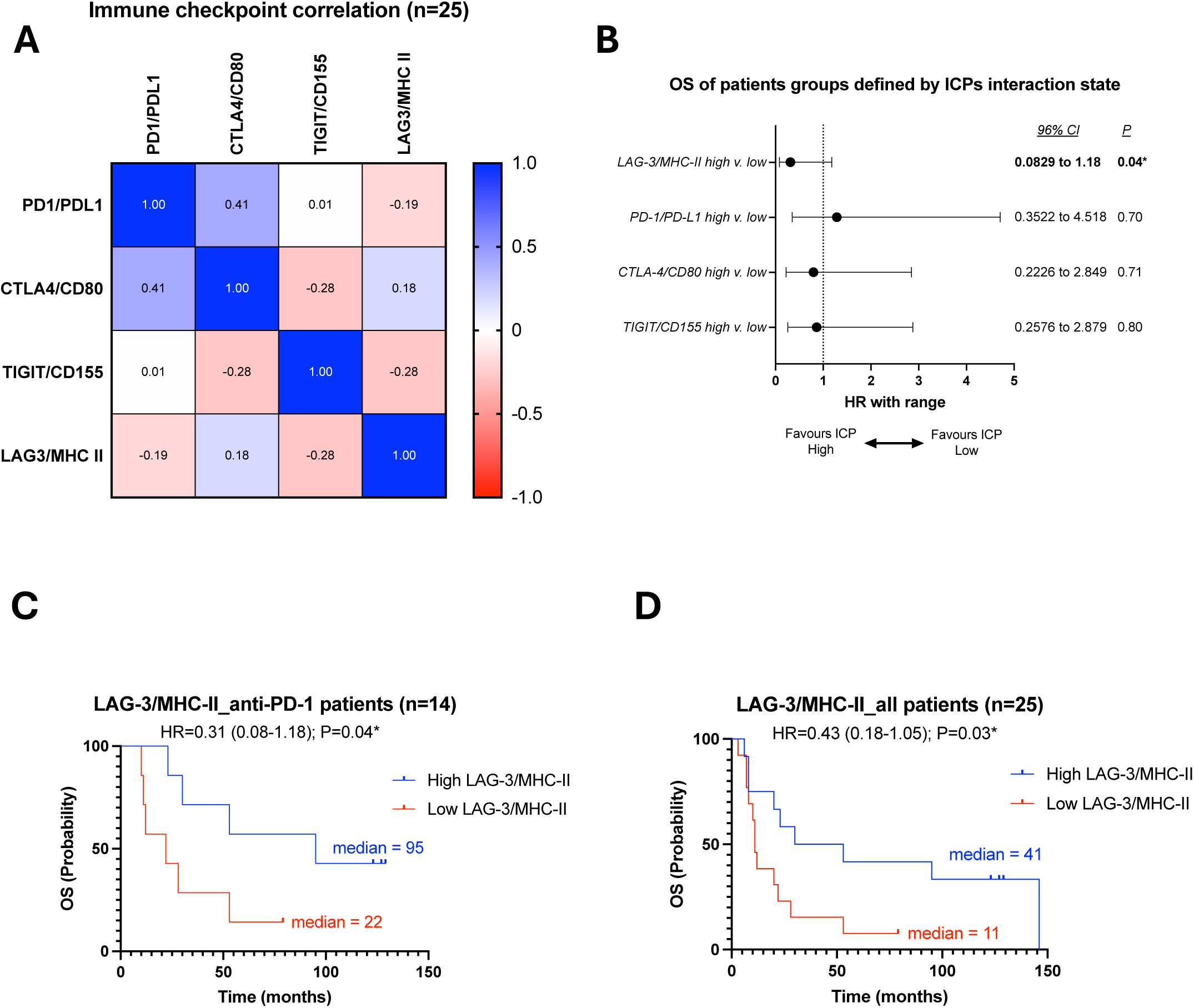
Overall survival of patients subgroup treated anti-PD-1-based therapies stratified by immune checkpoint interaction status. **(A)** Pearson correlation coeiicients (r; −1 to +1) are shown as a color-coded matrix, with red indicating negative correlations, blue indicating positive correlations, and colour intensity reflecting correlation strength. Only the correlation between PD-1/PD-L1 and CTLA-4/CD80 shows significance (r = 0.41; *P* = 0.04*). **(B)** Forest plot analysis of OS in patients separated by high vs. low immune checkpoint interaction. Hasard ratio with 95% confidence interval and *P* values are presented on the table for each ICP interaction **(C)** Kaplan–Meier analyses of melanoma patients treated with anti-PD-1-based therapies and stratified by high versus low immune checkpoint interaction for LAG-3/MHC-II. OS is favoured by LAG-3/MHC-II high interaction (*P* = 0.04*). **(D)** Kaplan–Meier analyses of melanoma patients treated with various immune checkpoint inhibitors and stratified by high versus low immune checkpoint interaction for LAG-3/MHC-II. OS is favoured by LAG-3/MHC-II high interaction (*P* = 0.03*).

### LAG-3/MHC-II engagement informs on immune checkpoint inhibitor response in the melanoma cohort

Exploiting the simultaneous evaluation of four ICP interactions within this limited cohort, we compared the potential of each ICP to predict response to ICI treatment. To this end, OS of patients stratified into high-versus low ICP-interaction groups were examined (Fig. 3B). The QF-Pro® scores cutoff values were determined using the online tool “Cutoff finder” [14] (Supplementary Table 3 and Materials and Methods). In this initial analysis patients treated with anti-CTLA4 monotherapy were excluded as 87% of the patients did not respond to treatment (Supplementary Table 4); CTLA-4 monotherapy showed a much lower median survival than PD-1 monotherapy or combination anti-PD-1 plus anti-CTLA-4 (8 vs. 41 or 49 months respectively; log rank comparison of the 3 curves *P* = 0.03; Supplementary Fig. 3).

The evaluation of the association between high vs. low interaction states and survival outcomes, was performed using a Cox proportional hazards analysis. The resulting hazard ratios (HRs) and 95% confidence intervals (CIs) are summarized in a forest plot (Fig. 3B), illustrating the effect size and direction across all immune checkpoints. Interestingly, despite the limited cohort size, LAG-3/MHC-II engagement demonstrated a significant association with improved survival, with an HR of 0.31 indicating an approximately 70% reduction in the risk of an event at any given time (i.e. favourable OS) with good significance and a tight 95% confidence interval in patients with high interaction levels (HR = 0.31, 95% CI 0.08–1.18; *P =* 0.04). In contrast, CTLA-4/CD80, TIGIT/CD155 and PD-1/PD-L1 interaction states did not display a consistent directional association with OS, as reflected by the wide 95% confidence interval. The Kaplan-Meier analysis of survival illustrates the association of LAG-3/MHC-II interaction state with the subgroup of patients who responded to anti-PD-1-based treatments (Fig. 3C). A high LAG-3/MHC-II interaction was associated with a marked OS benefit compared with low interaction levels (95 vs. 22 months; *P* = 0.04). Incorporating patients treated with ipilimumab (n = 25) into the survival analysis showed that a high LAG-3/MHC-II interaction state continued to identify patients with longer overall survival; this association was therefore independent of the immune checkpoint inhibitor used (41 vs. 11 months; *P* = 0.03; Fig. 3D).

## Discussion

Immune checkpoint inhibitors have transformed the therapeutic landscape of advanced malignancies; however, a substantial proportion of patients fail to respond, and the PD-L1 (TPS) expression biomarker currently used in clinical practice has shown limited predictive accuracy. This likely stems from its inability to directly inform on functional PD-1/PD-L1 engagement, making it a poor surrogate of the true immune checkpoint inhibitor target status. Conversely, the remarkable predictive value of PD-1/PD-L1 engagement for response to immunotherapy as compared to PD-L1 TPS in NSCLC [11], highlights the importance of exploiting such functional biomarkers to predict patient response to ICIs. Here, we extended previous findings to the analysis of the interaction states of four immune checkpoints: PD-1/PD-L1, CTLA-4/CD80, TIGIT/CD155, and LAG-3/MHC-II, all of which are considered to play a role in tumour immune evasion. Spatial localisation and quantification of these interactions across various settings enabled the assessment of the patterns of multiple engagements in situ, not reported previously.

Enhanced FRET–FLIM–based technology was employed to quantitatively measure these interactions (QF-Pro® scores) and to spatially resolve them (QF-Pro® maps) from cell models to tissue and tumour sections, providing a novel approach to interrogating the tumour immune landscape in FFPE samples. Consistent with prior observations in NSCLC, the sensitivity and specificity of the method enabled the detection of both intra- and inter-patient heterogeneity in checkpoint interactions. Moreover, differences observed between the spatial localisation of ICP interactions (QF-Pro® maps), and the corresponding receptor–ligand expression patterns, support the assertion that expression-based analyses do not reliably capture functional immune states. These observations are consistent with previous reports showing that direct quantification of checkpoint engagement provides a more informative indicator of tumour immune status [11]. These findings suggest that spatially resolved interaction mapping could provide an additional layer of information beyond interaction quantifications (QF-Pro® scores) alone, offering further functional insight into the TME ICP landscape.

Having validated QF-Pro® for the identification of ICP interactions in FFPE tumour tissues, a small cohort of melanoma patients treated with immune checkpoint blockade was used to enable simultaneous analysis of four ICPs per patient and to explore potential associations between their interaction states. Pearsons’ correlation analysis performed on this cohort showed a moderate but significant linear correlation (r = 0.41; *P* = 0.04) between PD-1/PD-L1 and CTL4/CD80 interactions while no correlated or exclusive distributions were found in any of the other ICP pairs. The correlation reflecting the co-activation of the two ICP pathways may suggest the co-infiltration of various immune cell types in the TME of several patients samples; i.e. CTLA-4 expressing cells primarily involved in early T-cell priming as well as exhausted cytotoxic T-cell expressing PD-1. Although the validation of this finding would require further investigation in a larger cohort, this analysis offers a first proof-of-concept for exploring how these pathways may co-occur and indeed where they function independently. The specific identification of patients in whom both pathways are highly active may improve the selection of responders to combination therapy while reducing the risk of immune-related adverse events (irAEs) in patients unlikely to benefit from this more aggressive treatment approach. Alternatively, as this co-regulation may reflect an activated yet autoinhibited immune state within the TME, these tumours may also remain sensitive to reactivation through anti–PD-1 monotherapy, as was recently showed in NSCLC [15].

Further analysis of this relatively small cohort was performed initially to evaluate whether any of the immune checkpoints interaction states could inform on patient survival following treaments that included PD-1 blockade. The patients were divided in two groups on the basis of high vs. low ICPs interaction states (QF-Pro® scores), identified using the comprehensive web-based application Cutoff finder [14]. The Cox proportional hazards and the Kaplan-Meier analysis of OS, uniquely identified patients with high LAG-3/MHC-II interaction state as faring well in these anti-PD-1-based treatments with a significant 73 months survival difference between the two groups (*P* = 0.04). Notably, extending the analysis to include CTLA-4 blockade indicated that this was a broad prognostic biomarker and not specific to anti-PD-1 treatment.

The mechanism underlying the association between LAG-3/MHC-II interaction and improved outcomes with anti–PD-1-based therapy may be explained by the fact that it could reflect a “hot” TME with the presence of inhibited but tumour-reactive T cells. In accordance, LAG-3 expression has been associated with CD8^+^ T-cell infiltration and an inflamed tumour microenvironment in melanoma, and its co-expression with PD-1 marks a therapeutically targetable exhausted T-cell population [16]. Accordingly, QF-Pro® map enabled the spatial identification of LAG-3/MHC-II positive immune cell infiltrations in melanoma (Fig.2D) as well as PD-1/PD-L1 positive infiltration in examples of melanoma and NSCLC (Fig.1B). Presently, single-agent anti–PD-1 therapy (pembrolizumab or nivolumab) typically remains the standard of care achieving objective response rates of about 40%. However, LAG-3 combination therapies (nivolumab plus relatlimab) have been approved for advance melanoma owing to greater efficacy compared to nivolumab monotherapy (RELATIVITY-047 [17] RELATIVITY-098 [18]), despite being associated with increased toxicity, with higher incidence of immune-related adverse events (irAEs) [18]. In RELATIVITY-047 clinical study, LAG-3 had been clearly established as a prognostic marker with high expression correlated to better prognosis.

Although recent clinical data suggest that LAG-3 expression alone does not reliably predict which patients will benefit from anti-LAG-3 therapy [17], quantification of LAG-3/MHC-II interactions, providing a more direct and robust measure of LAG-3 axis activation, may represent a more informative approach. This hypothesis would merit evaluation in cohorts of patients treated with nivolumab monotherapy or in combination with relatlimab.

While anti-TIGIT-targeted therapies are not yet clinically available, analysing TIGIT/CD155 interaction may offer important insights for drug development and therapeutic application. Although early results with anti-TIGIT agents, such as tiragolumab, generated significant optimism, outcomes from phase II and III trials have so far been disappointing [19]. As a result, patient stratification has been proposed as a strategy to improve therapeutic precision [20]. In this context, directly assessing TIGIT/CD155 interactions, more accurately reflecting immune checkpoint engagement, could be valuable for identifying patients most likely to benefit from such treatments.

Taken together, these findings underscore the complexity of immune regulation in cancer patients and support the rationale of parallel assessment of multiple immune checkpoints to better characterize the immune landscape. By capturing the concomitant or mutually exclusive regulation of immune checkpoint pathways, such an approach will provide a deeper and more accurate understanding of the mechanisms tumours employ to evade immune surveillance. This, in turn, may create opportunities for the development of more personalized immunotherapy strategies, offering a framework to optimize treatment selection, overcome resistance, and potentially reduce adverse effects. Validation of these approaches in larger cohorts will determine how well these biomarkers have predictive value in addition to prognostic significance (LAG-3 engagement).

## Authors contribution

All authors listed have made a substantial contribution to the work and approved it for publication. C.C.N and L.C. designed and performed the QF-Pro® experiments on FFPE samples. J.G., M.R. and J.M. designed and performed QF-Pro® experiments on cells assays. C.E.A and S.M.A carried out patient recruitment and provided the FFPE tumour samples and associated clinical data. J.A., B.B., J.M. and F.A. were involved in the development of the QF-Pro® FLIM platform. V.C. performed the various statistical analyses of the FRET (QF-Pro®) data with the clinical outcomes. C.E.A and S.M.A and V.C. were involved in the critical discussion of the clinical results. C.C.N, L.C. and V.C. were involved in the writing of the manuscript. P.J.P, J.G. and J.M. were involved in the critical reading of the manuscript. All authors read and approved the final manuscript.

## Data Availability

All data produced in the present work are contained in the manuscript

## Acknowledgments

This work was supported in part by the PREDICTIM project grant number CPP2021-008390.

## Ethics statement

The melanoma samples use are part of a study that was approved by the ethics committee of the University of Navarra, Spain (protocol 111/2010). FFPE tissue samples were obtained with informed patients’ consent and retrieved from the pathology files of the Clínica Universidad de Navarra.

## Materials and methods

### Primary antibodies

The primary antibody list is presented in Supplementary table 1.

### Secondary reagents

QF-Pro® reagent kits (HAWK Biosystems) contain all reagents needed to perform the labelling process. Briefly, the reagents provided within the kit include peroxidase suppressor, blocking and washing buffers, fluorescent (ATTO488 and AlexaFluor594) labelled secondary anti-mouse and anti-rabbit antibodies, amplification reagents and mounting medium.

### FFPE-tissue and tumour samples

Formalin-fixed, paraffin-embedded (FFPE) sections of 3-5 mm thickness were obtained from diagnostic biopsies, tumour resections or healthy tissues. Melanoma samples were provided by the Pathology Department of Clínica Universidad de Navarra (CUN). FFPE tissue samples were obtained with informed patients’ consent and retrieved from the pathology files of the Clínica Universidad de Navarra (protocol 111/2010). Tonsil blocks and TMA slides of breast tumours were purchased from Amsbio. FFPE samples were stained with H&E following standard protocol.

### QF-Pro® (Quantifying Function in Proteins) Assay

Before incubation with QF-Pro® reagents, pre-treatment processes of deparaffination, rehydration and epitope retrieval were performed using a Dako PT-Link. Samples were then treated following the QF-Pro® labelling protocol. Briefly, samples were incubated with endogenous peroxidase suppressor and then with blocking buffer to prevent non-specific signal. Samples were incubated with primary antibodies overnight at 4°C and then with donor and acceptor secondary reagents at room temperature (RT) for two hours. Amplification reagent was applied to Donor-Acceptor slides for 20 minutes and mounting medium was applied. Slides were stored until ready for fluorescence lifetime imaging microscopy analysis. A detailed protocol for QF-Pro® assays can be found in our previous publications [11, 21].

### Acquisition, analysis and quantification

Imaging and quantification of the immune checkpoint interaction states were performed using Violet 3.0. (Fluorescence Lifetime Imaging Microscope), controlled by QF-Pro® software (HAWK Biosystems). Violet 3.0 and QF-Pro® software automatically measure the decay of the lifetime of the donor fluorophore in the absence and presence of the acceptor (FRET-FLIM) to calculate interaction states (QF-Pro® scores). Violet 3.0 and QF-Pro® software allow for the spatial quantification of proteomic functional events across the sample. A decrease in donor lifetime (1D) in the presence of the acceptor chromophore (1DA) is indicative of resonance energy transfer. FRET efficiency (Eg % or QF-Pro® score) values were calculated using the following equation, where 1D and 1DA are the lifetimes of the donor in the absence and presence of the acceptor, respectively:

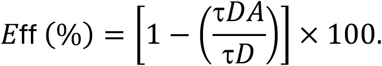

### Detailed Statistical analysis

Box-and-whisker plots were generated using GraphPad Prism 10. Here, the boxes represent the 25-75% range of the data and the whiskers represent the minimum and maximum values. Melanoma patients were ranked in order of their mean FRET efficiencies (QF-Pro® scores i.e. interaction status) and split into two groups using the webtool, Cutoff Finder, to define objectively a cut-og point for survival analysis. The tool, described by Budczies *et al.*, 2012, utilises R scripts [14]. The log-rank (Mantel–Cox) test was carried out to determine significant differences between the groups. Kaplan-Meier curves were then plotted using GraphPad Prism 10. The hazard ratio and P values were also calculated using GraphPad Prism 10. Correlation analyses were performed using the Pearson correlation test on continuous variables, and corresponding r values and two-tailed P values were generated automatically by the software. Correlation matrix was exported directly from Prism 10 for presentation. Statistical significance was defined as *P* < 0.05. Forest plot analyses were performed using GraphPad Prism 10 to visualize hazard ratios (HR) and corresponding 95% confidence intervals for each variables. Individual HR estimates were plotted as point estimates with horizontal confidence interval bars. A reference HR = 1 was used to indicate the null effect. Statistical calculations and graphical outputs were generated directly within Prism 10.

### Cell cultures and sensitivity assays

The commercially validated Promega Blockade Bioassay kits, originally designed to measure the antibody blockade of PD-1/PD-L1 and TIGIT/CD155 interaction by luminescence, were adapted for performing QF-Pro® protocol. Cells were thawed and directly used in this assay following the recommendations of the manufacturer. PD-L1 or CD155–expressing CHO-K1 cells were seeded onto 96 well plates and were incubated at 37 °C with 5% CO_2_ for 16 hours. PD-1 or TIGIT–expressing Jurkat cells together with the blocking antibodies (against PD-1 or TIGIT respectively) were added to the CHO-K1cells. The co-cultures were incubated for 20 hours at 37°C with 5% CO_2_. The unbound cells were removed by washing with PBS before being fixed with 4% PFA for 15 minutes. After fixation, cells were washed with PBS and stored at 4 °C until labelling. QF-Pro® labelling was performed as described for FFPE-samples.

## Supplementary figures legends

**Supplementary Figure 1: Quantifying Function in Proteins (QF-Pro®) technology**. The technology comprises specific fluorescent secondary reagents, Fluorescence Lifetime Imaging Microscopy (FLIM) platform and dedicated software package for quantification of protein complex formation or post translational modifications in cells or in FFPE tissue samples. The cartoon illustrates the interaction between two proteins recognised by primary and fluorescent secondary antibodies. The interaction is detected by quantifying the transfer or energy (FRET) between the 488nm-laser excited donor (ATTO488) and the amplified acceptor (Alexa Fluor 594). FRET only occurs only when the two proteins are at a binding distance (< 10 nm) and is rapidly lost when the distance between the proteins increases.

**Supplementary Figure 2: Melanoma cohort clinical data analysis**. **(A)** Forest plot presenting the impact of various clinical parameters on patients OS. **(B)** Box plot presenting the OS of patients with or without metastasis. Each dot represent at patient. **(C)** Proportion of patients presenting primary or metastatic tumours who show benefit at 12 months.

**Supplementary Figure 3. Kaplan-Meier analysis of the melanoma patients survival dependent upon immune checkpoint inhibitors treatment.** Median survival for anti-CTLA4 compared with anti-PD-1 monotherapy (8 vs. 41 months; *P* = 0.02*) and anti-CTLA4 with anti-PD-1 plus anti-CTLA-4 combination (8 vs. 49 months; *P* = 0.006**). The table presents the survival proportion and the number at risk in brackets, over a period of 137 months.

## References

1. Abdeladhim, M., J.L. Karnell, and S.A. Rieder, In or out of control: Modulating regulatory T cell homeostasis and function with immune checkpoint pathways. Front Immunol, 2022. 13: p. 1033705.

2. Galassi, C., et al., The hallmarks of cancer immune evasion. Cancer Cell, 2024. 42(11): p. 1825–1863.

3. He, X. and C. Xu, Immune checkpoint signaling and cancer immunotherapy. Cell Res, 2020. 30(8): p. 660–669.

4. Kong, X., et al., Immune checkpoint inhibitors: breakthroughs in cancer treatment. Cancer Biol Med, 2024. 21(6): p. 451–72.

5. Berland, L., et al., Further knowledge and developments in resistance mechanisms to immune checkpoint inhibitors. Front Immunol, 2024. 15: p. 1384121.

6. Iranzo, P., et al., Overview of Checkpoint Inhibitors Mechanism of Action: Role of Immune-Related Adverse Events and Their Treatment on Progression of Underlying Cancer. Front Med (Lausanne), 2022. 9: p. 875974.

7. Maloney, A.K., et al., Nivolumab maintenance improves overall survival of patients with advanced melanoma who experience severe immune-related adverse events on nivolumab plus ipilimumab. J Immunother Cancer, 2024. 12(8).

8. Kreft, S. and P. Lorigan, Combination Immunotherapy for Advanced Melanoma-How to Choose? J Clin Oncol, 2025. 43(4): p. 478–479.

9. Twomey, J.D. and B. Zhang, Cancer Immunotherapy Update: FDA-Approved Checkpoint Inhibitors and Companion Diagnostics. AAPS J, 2021. 23(2): p. 39.

10. Yamaguchi, H., et al., Advances and prospects of biomarkers for immune checkpoint inhibitors. Cell Rep Med, 2024. 5(7): p. 101621.

11. Sanchez-Magraner, L., et al., Functional Engagement of the PD-1/PD-L1 Complex But Not PD-L1 Expression Is Highly Predictive of Patient Response to Immunotherapy in Non-Small-Cell Lung Cancer. J Clin Oncol, 2023. 41(14): p. 2561–2570.

12. Massoni-Badosa, R., et al., An atlas of cells in the human tonsil. Immunity, 2024. 57(2): p. 379–399 e18.

13. Guo, C., et al., TIGIT as a Novel Prognostic Marker for Immune Infiltration in Invasive Breast Cancer. Comb Chem High Throughput Screen, 2023. 26(3): p. 639–651.

14. Budczies, J., et al., Cutoff Finder: a comprehensive and straightforward Web application enabling rapid biomarker cutoff optimization. PLoS One, 2012. 7(12): p. e51862.

15. Juan Gumuzio, J.M., Nicole Quimi et al., Immune Checkpoint Crosstalk: CTLA-4/CD80 Engagement as a Predictor of Anti-PD-1/PD-L1 Therapy Outcome in NSCLC. Research Square, 2025. 08 July 2025, **PREPRINT (Version 1)**(08 July 2025).

16. Vryza, P., I. Georgakopoulos-Soares, and A. Zaravinos, Predicting skin melanoma progression via LAG-3, TIGIT and HAVCR2. Funct Integr Genomics, 2026. 26(1).

17. Tawbi, H.A., et al., Relatlimab and Nivolumab versus Nivolumab in Untreated Advanced Melanoma. N Engl J Med, 2022. 386(1): p. 24–34.

18. Long, G.V., et al., Adjuvant nivolumab and relatlimab in stage III/IV melanoma: the randomized phase 3 RELATIVITY-098 trial. Nat Med, 2025. 31(12): p. 4301–4309.

19. Cho, B.C., et al., Tiragolumab plus atezolizumab versus placebo plus atezolizumab as a first-line treatment for PD-L1-selected non-small-cell lung cancer (CITYSCAPE): primary and follow-up analyses of a randomised, double-blind, phase 2 study. Lancet Oncol, 2022. 23(6): p. 781–792.

20. Sundstrom, E.C., X. Huang, and A.J. Wiemer, Anti-TIGIT therapies: a review of preclinical and clinical efficacy and mechanisms. Cancer Immunol Immunother, 2025. 74(8): p. 272.

21. Sanchez-Magraner, L., et al., High PD-1/PD-L1 Checkpoint Interaction Infers Tumor Selection and Therapeutic Sensitivity to Anti-PD-1/PD-L1 Treatment. Cancer Res, 2020. 80(19): p. 4244–4257.

