## Supplementary figures and images for "Simultaneous quantification of multiple immune checkpoint interactions in melanoma"

### Supplementary Figure 1

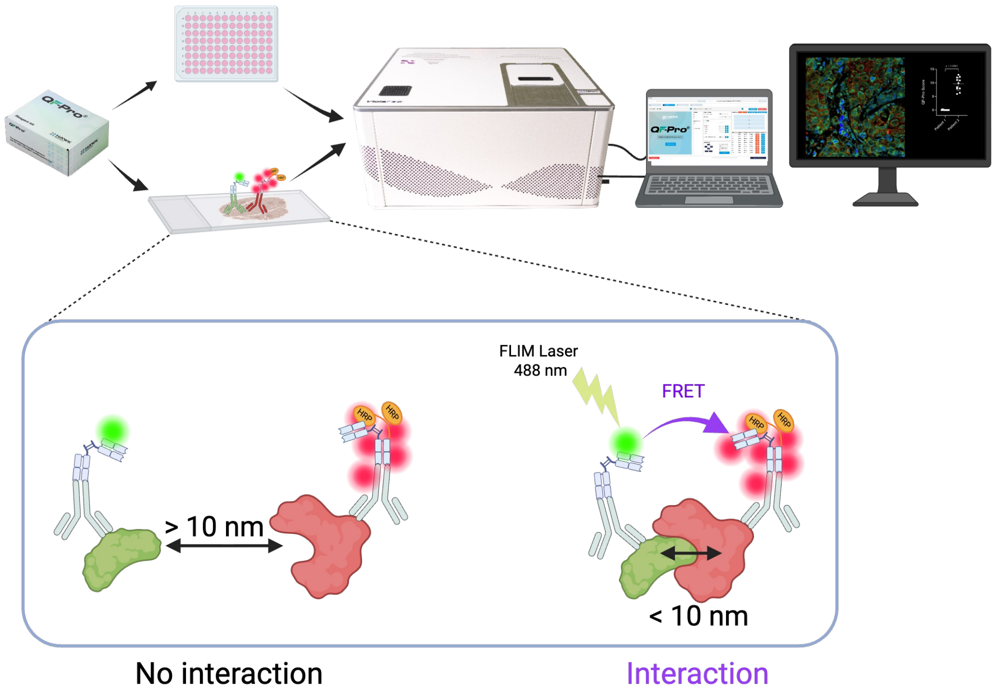


Supplementary Figure 1 Cacho et al.

### Supplementary Figure 2

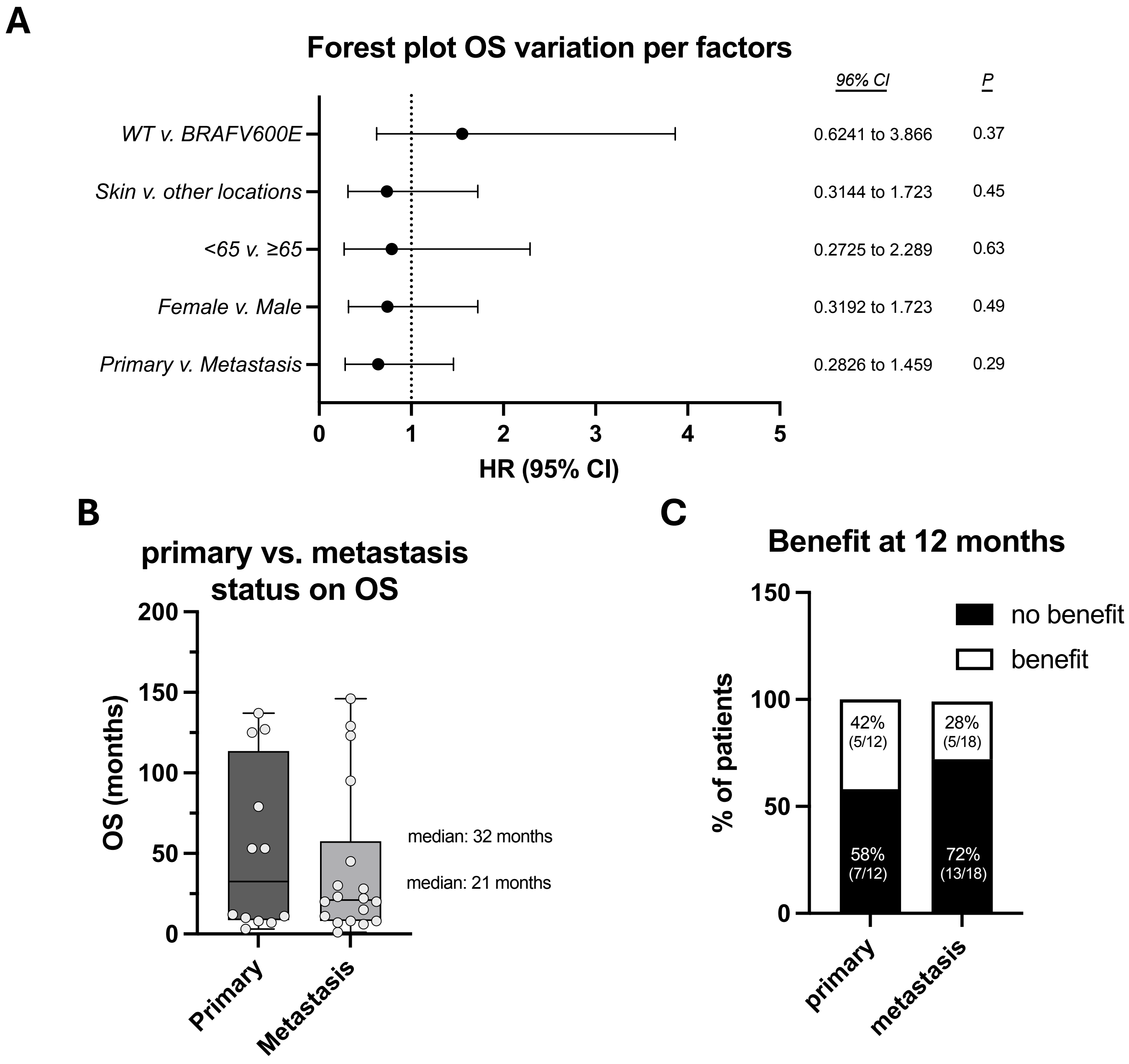


Supplementary Figure 2 Cacho et al.
