## Supplementary Figure 3 for "Simultaneous quantification of multiple immune checkpoint interactions in melanoma"

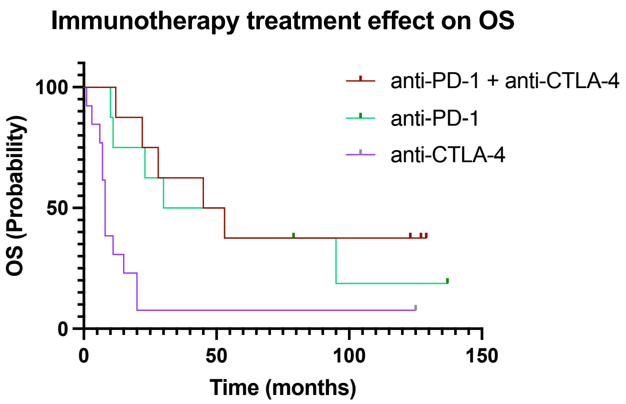


| Survival proportion and Number at risk | Time (months) | | | | |
|---|---|---|---|---|---|
| | **0** | **12** | **24** | **60** | **137** |
| anti-PD-1 | 100 (8) | 75 (7) | 62 (6) | 37 (4) | 19 (1) |
| anti-CTLA-4 | 100 (13) | 31 (5) | 7.7 (3) | 7.7 (3) | 7.7 (1) |
| anti-PD-1/anti-CTLA-4 | 100 (8) | 87 (8) | 75 (7) | 37 (4) | 37 (1) |

Supplementary Figure 3 Cacho *et al.*
