## Supplementary Table 1 for "Simultaneous quantification of multiple immune checkpoint interactions in melanoma"

**Supplementary table 1: Primary antibody selection.**

| Antibody | Catalogue Number |
|---|---|
| Anti-PD-1 NAT105 mouse mAb (Recombinant) | ab234444 |
| Anti-PD-L1 28-8 rabbit mAb | ab205921 |
| Anti-TIGIT 4A10 mouse mAb | RF16054 |
| Anti-CD155 rabbit mAb | NBP2-78711 |
| Anti-LAG3 mouse mAb | ab307517 |
| Anti-MHC II rabbit mAb | ab92511 |
| Anti-CTLA4 mouse mAb | ABIN6992223 |
| Anti-CD80 rabbit polyclonal Ab | MBS2522916 |
| TIGIT blocking antibody | Promega J2051 |
| PD-1 blocking antibody | Promega J1201 |
