## Supplementary Table 2 for "Simultaneous quantification of multiple immune checkpoint interactions in melanoma"

**Supplementary table 2: General clinical data of the melanoma cohort.**

| | **Patient cohort (n=30)** | **Information** |
|---|---|---|
| male/female | 18/10 | • 2 patients information missing |
| BRAF V600E / wild type | 8 (27%)/20 | • 2 patients information missing<br>• 6 patients treated with BRAF inhibitor ±MEK inhibitor before ICI |
| location skin/other location | 17/13 | - |
| Metastatic patients/non metastatic | 18 (58%)/12 | - |
| Age>65/<65 and (median age) | 23/7 (52) | - |
