## Supplementary Table 3 for "Simultaneous quantification of multiple immune checkpoint interactions in melanoma"

**Supplementary table 3: Immune checkpoints interactions cutoffs values**

| **Immune checkpoint interactions** | **QF-Pro® scores (%) cutoffs – high vs low interactions.** |
|---|---|
| PD-1/PD-L1 | High >1.35%> Low |
| CTLA-4/CD80 | High >1.25%> Low |
| TIGIT/CD155 | High >2.41%> Low |
| LAG-3/MHC-II | High >0.96%> Low |
