## Supplementary Table 4 for "Simultaneous quantification of multiple immune checkpoint interactions in melanoma"

**Supplementary table 4: Melanoma cohort clinical data analysis per treatment.**

| | anti-PD-1 | anti-CTLA-4 | anti-PD-1/CTLA-4 combination |
|---|---|---|---|
| Benefit at 12 months- patients with benefit/total (%) | n = 3/8 (37%) | n = 2/13 (13%) | n = 5/8 (62%) |
| Median DoR (months) | 11 | 4 | 19 |
| Median OS (months) | 41 | 8 | 49 |
| Metastatic patients/total (%) | n = 3/8 (37%) | n = 10/14 (71%) | n = 5/8 (62%) |
